# RETROSPECTIVE STUDY OF PATTERNS OF FAILURE IN CUTANEOUS SQUAMOUS CELL CARCINOMA TREATED WITH PRIMARY SURGERY: A TATA MEDICAL CENTER EXPERIENCE

**DOI:** 10.64898/2026.07.02.26357153

**Authors:** Pratiksha Tyagi, Santam Chakraborty, Anand Bardia, Karnav Panchal, Amrita Kaur, Sougata Maity, Gautam Biswas

## Abstract

**Background:** Cutaneous squamous cell carcinoma (cSCC) accounts for a significant proportion of skin malignancies in India, yet data on patterns of failure, particularly for extremity and truncal primaries remain scarce. We audited a decade of surgically treated cSCC at a tertiary cancer center to characterize failure patterns and associated risk factors.

**Methods:** This retrospective study included 161 patients with histopathologically confirmed cSCC treated surgically between January 2013 and December 2023, comprising 127 upfront/residual and 34 recurrent presentations. Primary sites were extremities (64%), head and neck (26%) and torso (10%). 21 patients had Marjolin’s ulcer. Outcomes included local, regional and distant failure, recurrence-free survival and overall survival. Brigham and Women’s Hospital (BWH) staging was applied to assess prognostic utility. Statistical analysis was done using Kaplan-Meier and competing-risk methods.

**Results:** Median follow-up was 2.4 years. Regional recurrence was the predominant failure pattern seen in 26 patients, local recurrence was seen in 14 patients and distant metastasis in 13. The 3-year cumulative incidences of local, regional and distant failure were 11%, 19% and 8.4% respectively. Rates of regional recurrence were substantially higher than Western series. Extremity primaries accounted for 19/26 regional recurrences. BWH T2b disease showed the highest regional failure rate (27.6%), exceeding T3 (17.8%) and T2a (6%) with perineural invasion significantly associated with regional failure in T2b/T3 tumors (p<0.001). Median time to regional metastasis was 8.4 months. At 3 years, overall survival was 77% and progression-free survival was 64%.

**Conclusion:** Regional recurrence is the dominant mode of failure in this cohort, at rates higher than most published series, with extremity primaries and BWH T2b staging identifying particularly high-risk subgroups. These findings highlight the need for a comprehensive staging system encompassing non head and neck cSCC and support prospective evaluation of elective nodal staging and adjuvant radiotherapy in high-risk patients, alongside intensified surveillance.

## INTRODUCTION

Cutaneous Squamous Cell Carcinoma (cSCC) is the second most common type of non-melanoma skin cancer (NMSC), accounting for approximately 20% of all skin cancers in the United States.(1) Among individuals with darker skin, cSCC is more prevalent than basal cell carcinoma.(2) In India, the reported incidence of skin cancer is relatively low, ranging between 0.5 and 4.8 per 100,000 in females and 0.04 to 6.2 per 100,000 in males.(3) As cancer is not a notifiable disease in low- and middle-income countries (LMICs) such as India, registry coverage remains incomplete, likely obscuring true incidence rates.

Lashiram et al. and Deo et al. reported cSCC accounting for 43.6% (40/92) and 55.8% (43/77) of all skin cancers in their respective case series from India.(4,5) These figures underscore that cSCC represents a significant proportion of skin malignancies in the Indian population, particularly in regions with distinct exposure patterns.

Failure patterns in cSCC typically include local recurrence at the primary site, regional nodal metastasis and less commonly, distant spread. These patterns are influenced by tumour-related factors including anatomical location, tumour size and depth of invasion, as well as host factors such as immune status.

Existing literature on recurrence patterns in cSCC predominantly focuses on head and neck primaries and sun-exposed sites. Notably, the American Joint Committee on Cancer (AJCC) 8th edition staging addresses cSCC of the head and neck separately, with no equivalent dedicated staging system for other anatomical sites such as the extremities and trunk. These less sun-exposed regions may have distinct mutational profiles and biological behaviours, warranting independent study.

A systematic search of PubMed using keywords pertaining to cSCC, recurrence patterns, and failure patterns restricted to India retrieved only a single relevant result (Teli et al., 2009; PMID: 20101335), reporting that regional nodal recurrence accounted for 21.72% (53/244) of failures in squamous cell cancer of trunk and lower extremities. (6) Given the paucity of data we audited the patterns of failure in our center over the past decade.

The objective of this study was to evaluate the patterns of failure and associated risk factors in patients with cSCC who underwent surgical treatment at our tertiary cancer center over a decade. To place these findings in context, key international series are briefly summarised. Soleymani et al. observed local recurrence in 22 (3.8%), regional recurrence in 32 (5.5%) and distant metastases in 11 (1.9%) of 527 patients with cSCC across various subsites.(7) Schmults et al., in a 10-year retrospective study of 985 patients with a median follow-up of 50 months, noted local recurrence in 45 (4.6%), regional nodal recurrence in 36 (3.7%) and cSCC-related death in 21 (2.1%).(8)

## MATERIALS AND METHODS

### Study Design and Patient Selection

A retrospective observational analysis was conducted at a tertiary cancer center in eastern India. Electronic hospital records and operative theatre records were searched to identify patients with histopathologically confirmed cSCC who underwent surgery between January 2013 and December 2023. Data were retrieved from the hospital’s management system database and supplemented by departmental logbooks of department of plastic surgery.

The study encompassed upfront cases, cases with residual disease and those presenting with recurrence. Patients with primary disease at any anatomical site were eligible, including those with Marjolin’s ulcer.

Variables collected included patient demographics, disease status at presentation and type of surgery performed. Histopathological parameters reviewed included maximum tumour size, lymphovascular invasion (LVI), perineural invasion (PNI), depth of invasion, skin layer involved, number of positive lymph nodes and total nodes retrieved in patients who underwent nodal dissection. Patients who failed to attend follow-up were contacted telephonically and encouraged to attend their scheduled appointment.

### Outcomes

The primary outcome was the patterns of failure. Secondary outcomes included recurrence-free survival (RFS) and overall survival (OS).

The events that were studied were local failure, regional failure, distant metastasis and death. Local failure was defined as the biopsy proven cSCC in the previously operated site. Regional failure was defined as biopsy/FNAC proven cSCC in the regional lymph nodal area. Distant metastasis was defined as radiological or histopathological evidence of cSCC in a region distant from the locoregional primary or regional nodal site(s). Patients who had nodal disease at initial presentation were analysed separately and were not included in the regional recurrence analysis.

Recurrence free survival was defined as the duration between the date of surgery to the duration of first progression or date. Patients who did not have any progression and were alive were censored. Overall survival was calculated from the date of surgery to the date of death due to any cause. Patients who were alive at the last follow up were censored. Cumulative incidence of regional failure was calculated as the time from date of surgery till the time of first regional failure.

### Treatment

Adjuvant radiotherapy (RT) was administered to patients with high-risk features like close or positive surgical margins, pathological nodal involvement or perineural invasion following multidisciplinary tumour board discussion. RT was most commonly delivered to a dose of 60 Gy in 30 fractions; alternative schedules of 55 Gy in 20 fractions and 50 Gy in 25 fractions were also used. All patients were routinely followed up in the outpatient departments of plastic surgery and radiation oncology.

### Statistical Analysis

For reporting of data, mean, median and Interquartile range (IQR) were used for numerical variables and frequency was calculated for categorical variables. Mean, range, median and IQR were calculated for numerical variables. Frequencies were reported for categorical variables. Time to event analysis was done using Kaplan Meier method. Cumulative incidence of local, regional and distant metastasis were calculated using the a competing risk model with death as the competing outcome.

## RESULTS

A total of 161 patients were identified. The most common primary sites were the extremities (n=103, 64%), head and neck (n=42, 26%) and torso (n=16, 10%). Twenty-one patients had Marjolin’s ulcer. Of the total cohort, 127 patients presented with primary or residual disease and 34 presented with recurrent disease. The median follow-up was 2.4 years.

Fifty-one patients (including 34 who presented with recurrence) had undergone their index surgery at an outside facility. Of these, 48/51 (94%) had undergone wide local excision (WLE); nodal dissection had been performed in only 2 patients. Slide and block review was performed for all patients referred from outside. Positive margins were identified in 12/51 (23.5%) and close margins in 10/51 (19.6%).

The remaining 110 patients underwent their primary surgery at our centre. Of these, 61/110 (55.5%) underwent WLE alone. Nodal dissection was performed in 45/110 (40.9%) patients simultaneously with WLE in 38 patients (84.4%) and as a staged procedure in 7 patients (15.6%). Overall, 36/161 patients (22.4%) had positive or close surgical margins, while 116/161 (72.0%) had clear margins.

In the adjuvant setting, 30/161 (18.6%) of patients received adjuvant radiotherapy. Of these, 16 patients received RT to tumor bed alone, 12 received to both tumor bed and draining nodal region and 2 to nodal basin alone. 11 patients had nodal recurrence (36.6%) of which 7 were seen in patients who received RT to tumor bed alone.

The demographic and clinicopathological characteristics of the cohort are presented in Table 1.

**Table 1:**
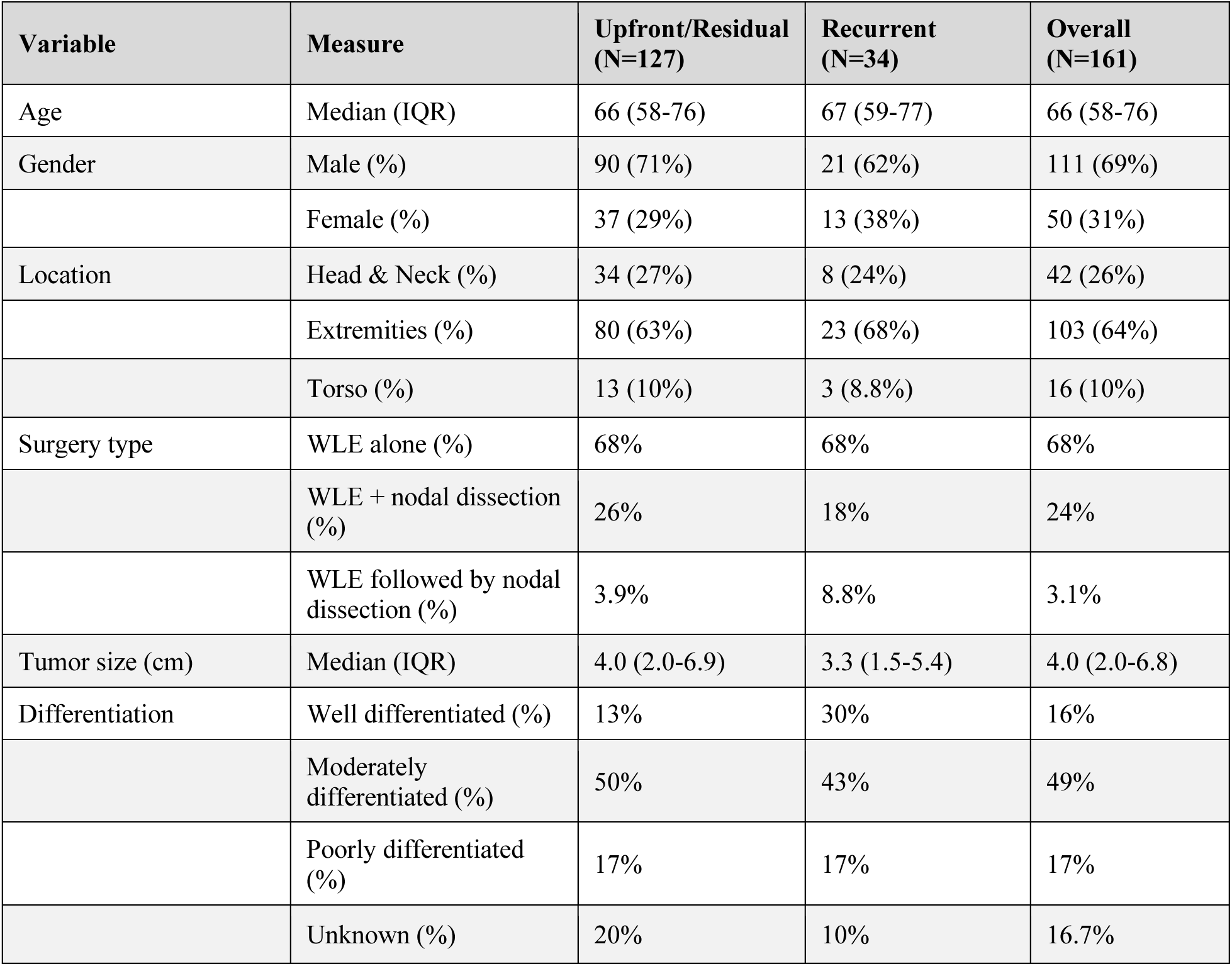
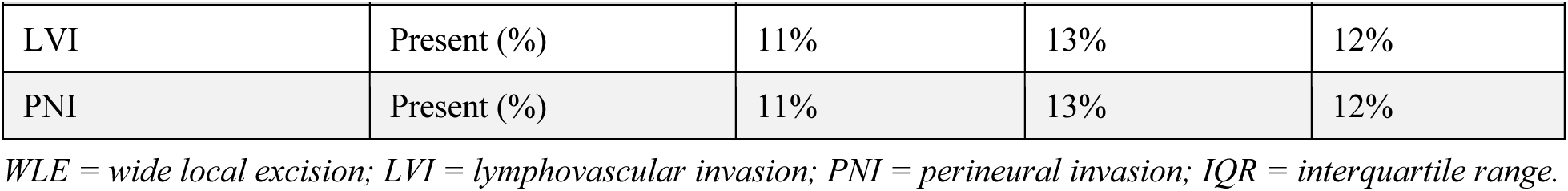
Clinicopathological characteristics of the study cohort.

### Overall Patterns of Failure

In the entire cohort, 26 patients had regional recurrence, 14 had local recurrence and 13 had distant failure, either in isolation or in combination. At 3 years, the overall survival was 77% (95% CI: 69%-86%) and the progression-free survival was 64% (95% CI: 56%-74%). At 6 years, OS was 67% and PFS was 56%, reflecting an approximate 10% absolute reduction over the subsequent 3 years.

The 3-year cumulative incidences of local recurrence, regional recurrence and distant metastasis were 11% (95% CI: 6.1%-18%), 19% (95% CI: 13%-27%), and 8.4% (95% CI: 4.4%-14%), respectively. Figure 1 depicts the overall failure pattern using an upset plot.

**Fig 1:**
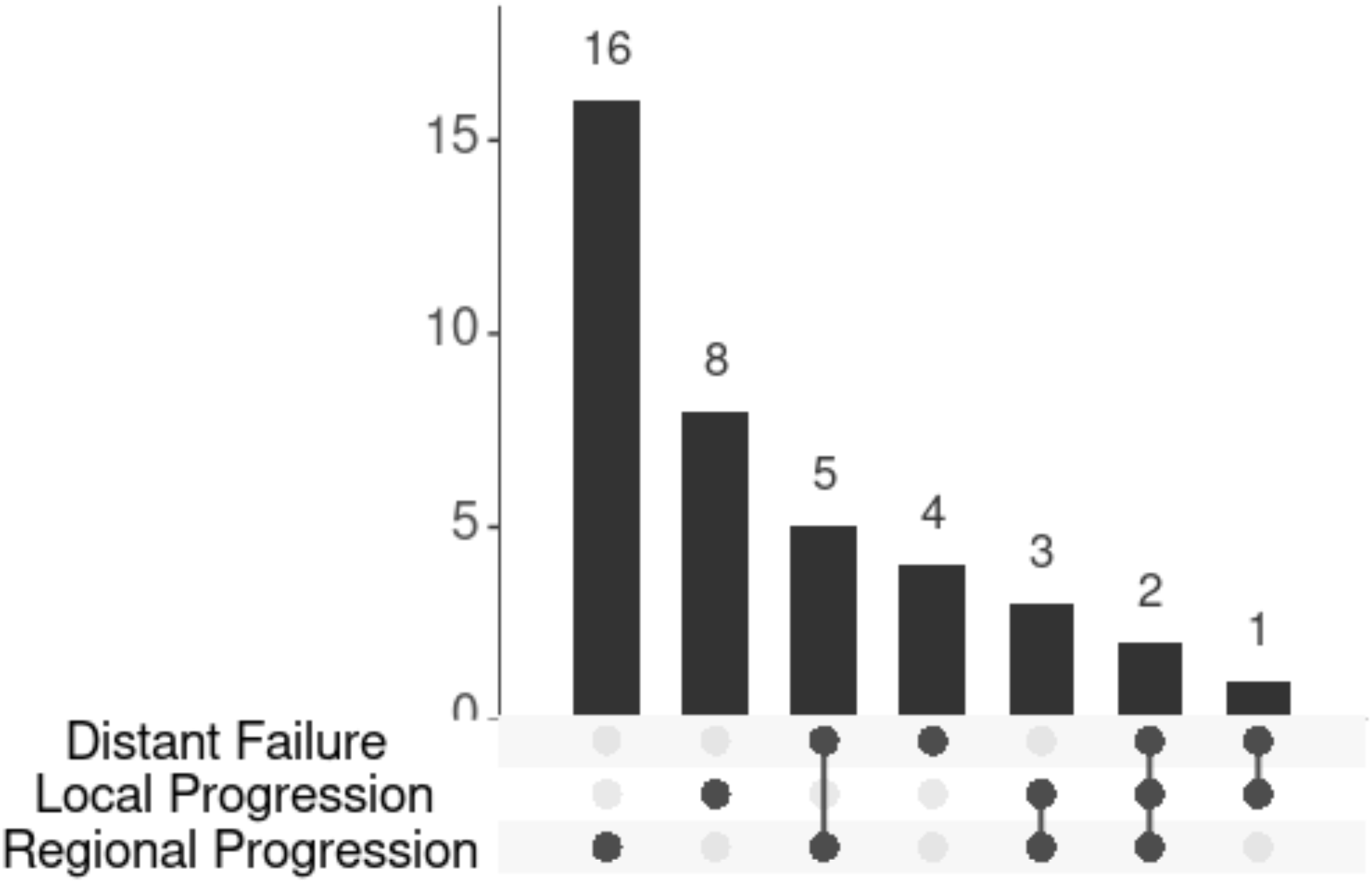
Upset plot showing patterns of failure across the entire cohort

### Patterns of Failure in Upfront/Residual Disease

Of the 26 total regional recurrences, 19 occurred in who were treated upfront (primary disease or residual disease). Among this subgroup, regional recurrences were observed in 4/34 (11.7%) patients with head and neck cSCC, 14/80 (17.5%) with extremity cSCC and 1/13 (7.6%) with torso cSCC (Fig 2).

**Fig 2:**
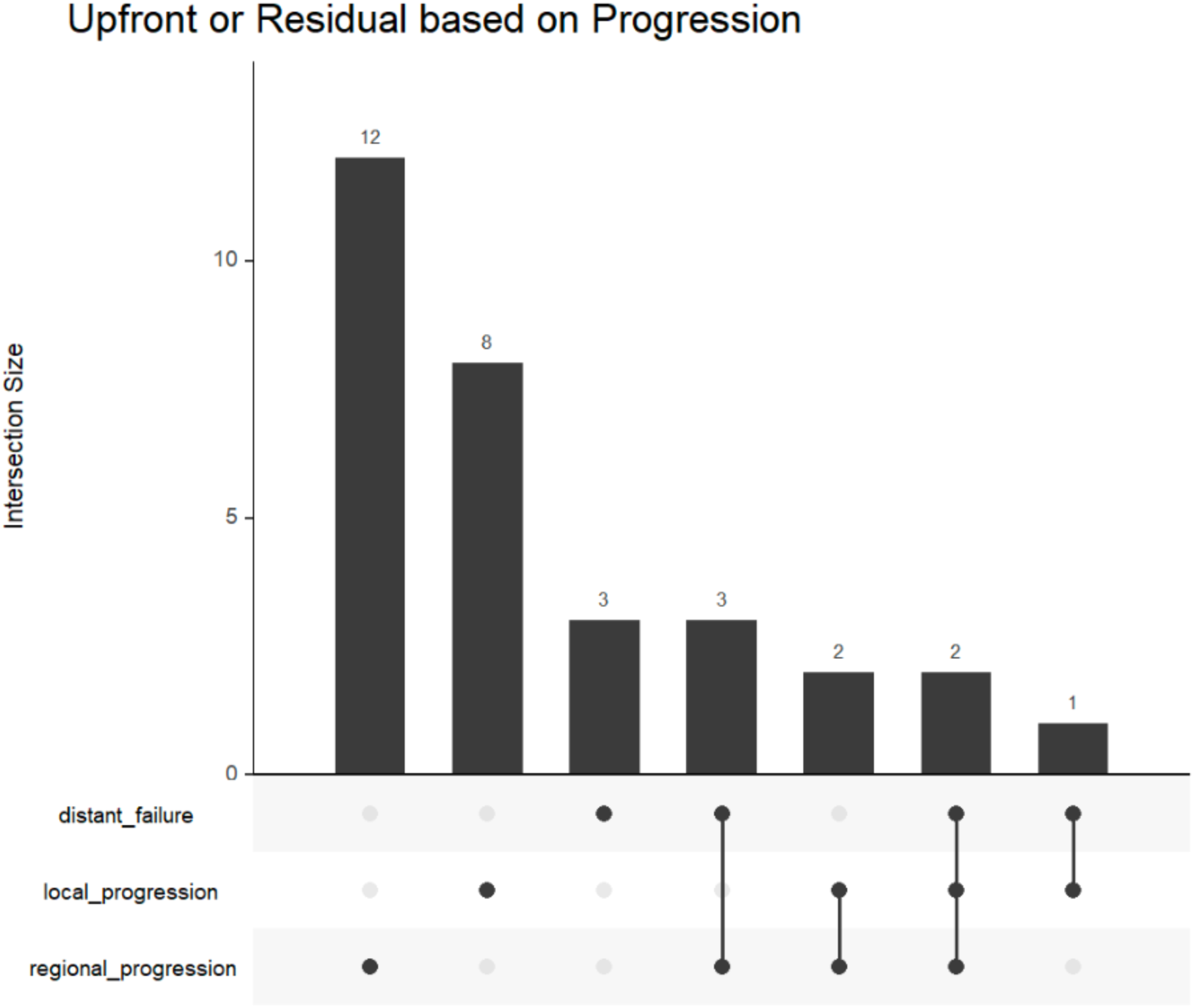
Upset plot showing patterns of failure in patients with upfront or residual disease

Regional recurrences occurred in 9/19 (47.3%) patients who had undergone WLE alone and in 7/19 (36.8%) who had undergone WLE with nodal staging surgery. Among patients who had not undergone nodal staging, 5/19 (26.3%) developed regional recurrence.

The 3-year cumulative incidences of local and regional recurrence in this subgroup were 13% (95% CI: 6.9%-21%) and 18% (95% CI: 11%-26%), respectively (Fig 4). The 3-year disease-free survival was 64% (95% CI: 55%-75%) and overall survival was 76%.

### Patterns of Failure in Recurrent Disease

Of the 26 total regional recurrences, 7 occurred in patients who had presented with recurrent disease at baseline. The 3-year PFS in this subgroup was 65% (95% CI: 50%-85%). The 3-year cumulative incidences of local and regional recurrence were 4.1% (95% CI: 0.27%-18%) and 24% (95% CI: 10%-42%), respectively (Fig 3, Fig 4).

**Fig 3:**
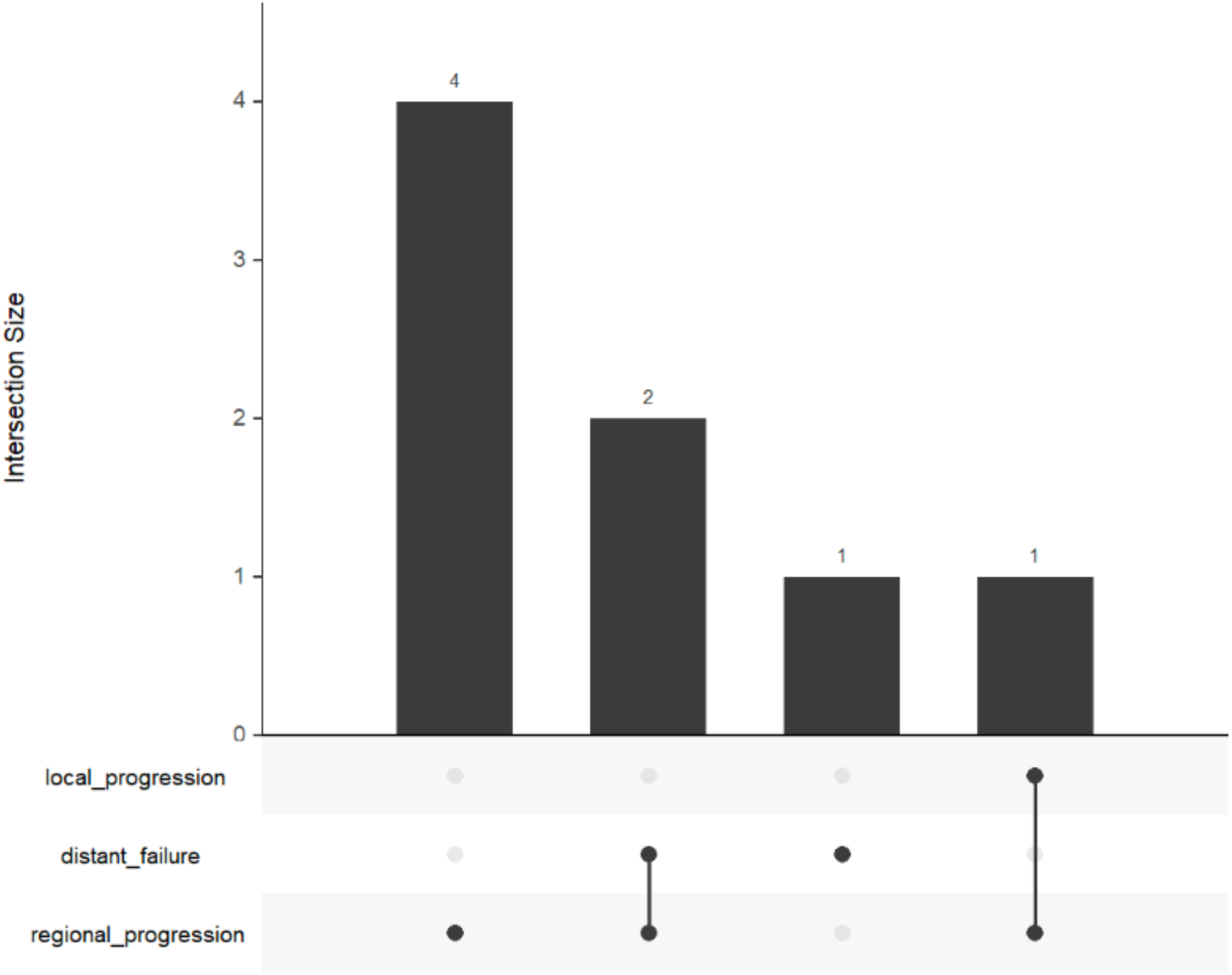
Upset plot showing patterns of failure in patients presenting with recurrent disease

**Fig 4:**
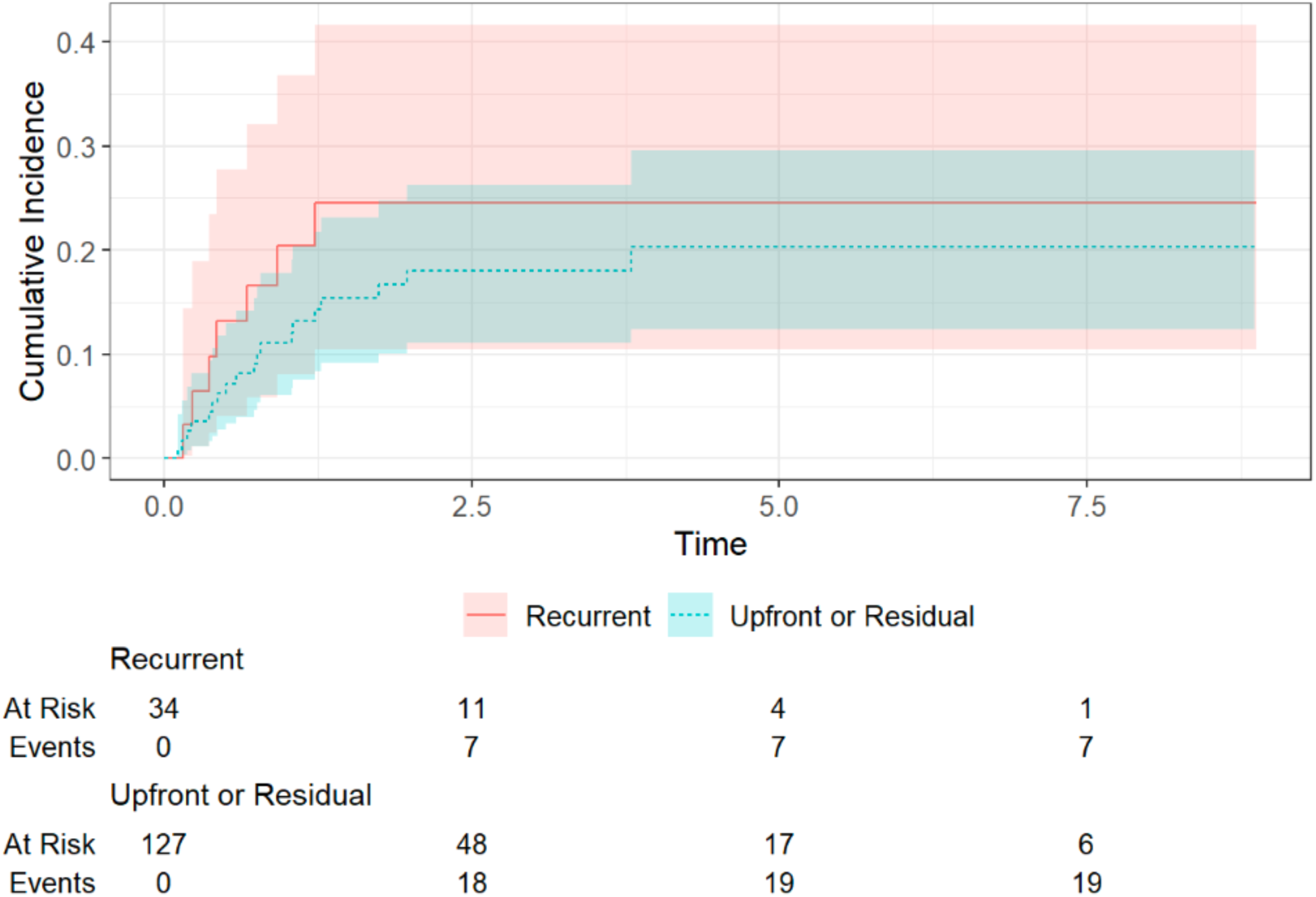
Cumulative incidence curves of regional recurrence stratified by disease presentation: competing risks model

**Fig 5:**
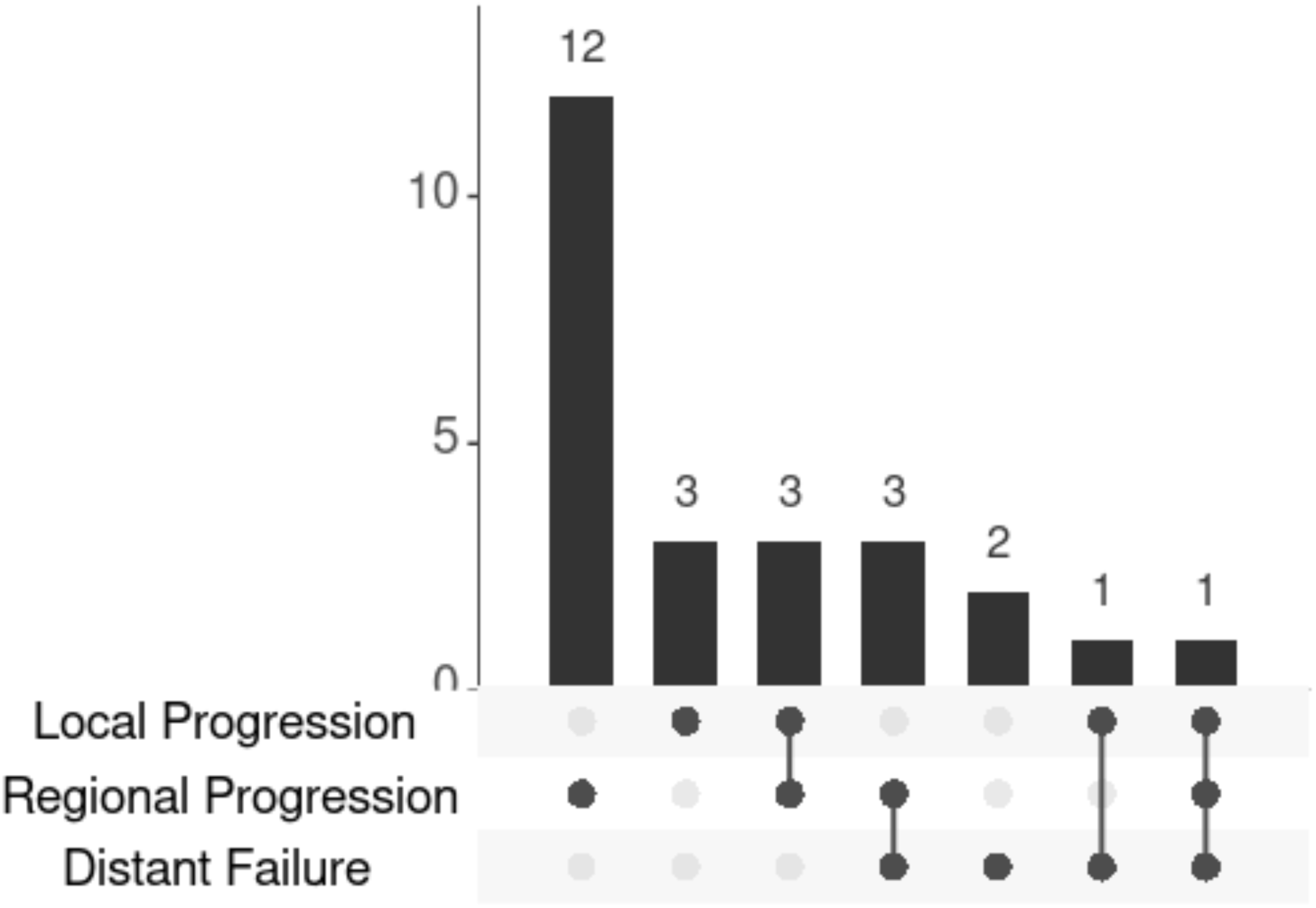
Upset plot showing patterns of failure in cSCC of the extremities

**Fig 6:**
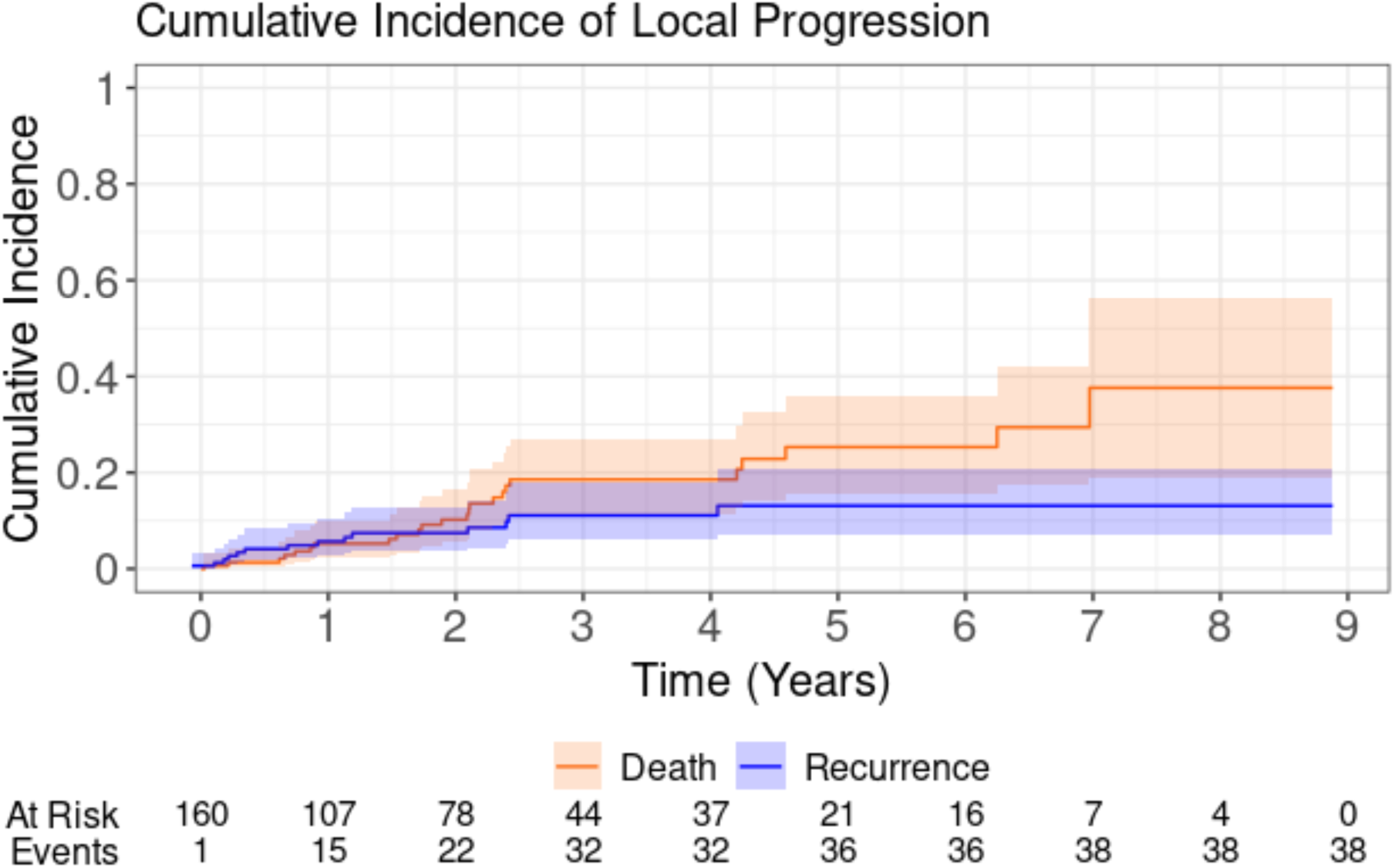
Kaplan–Meier curve showing cumulative incidence of local progression

**Fig 7:**
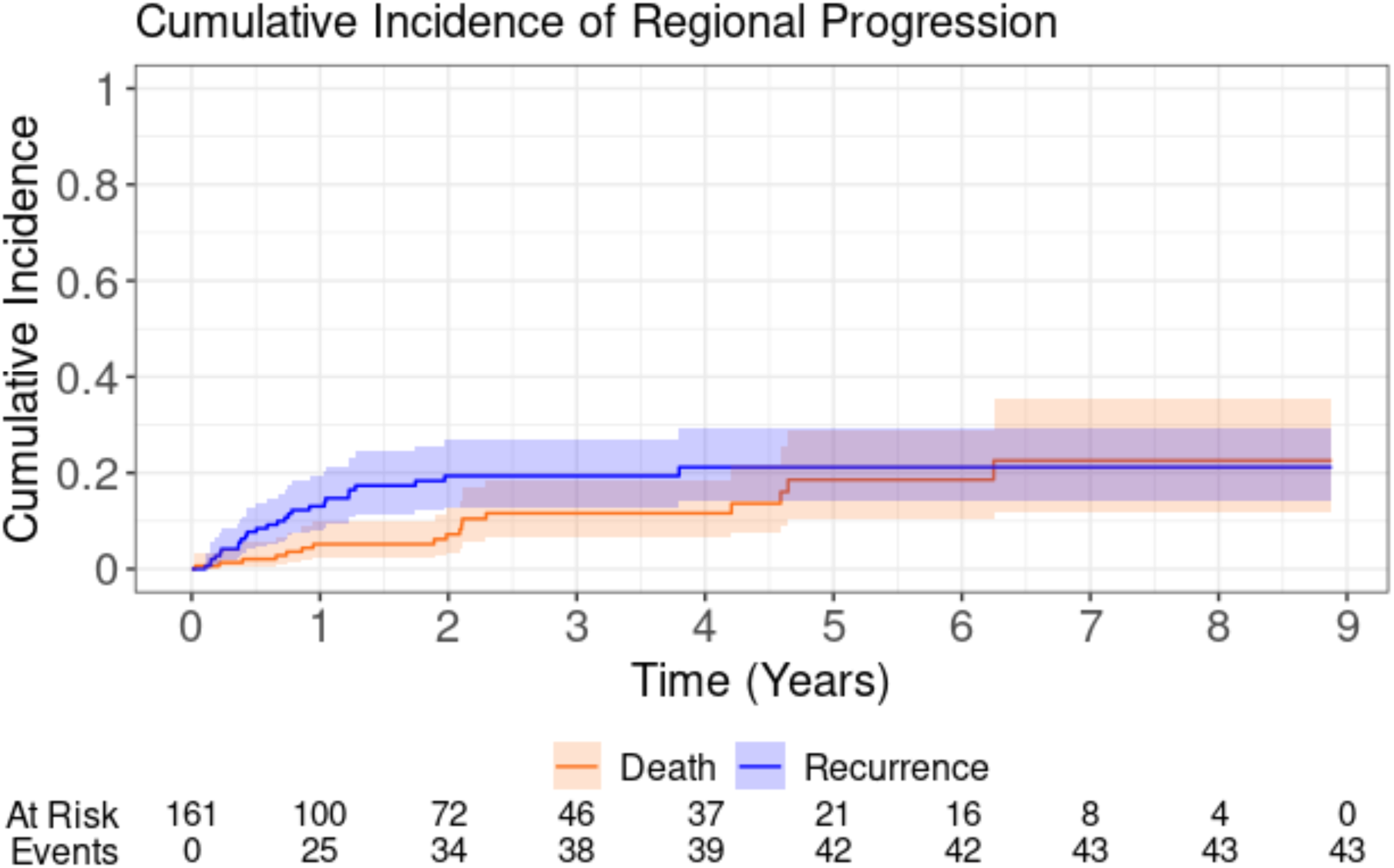
Kaplan–Meier curve showing cumulative incidence of regional progression

### Failure Patterns by Anatomic Site

Regional failure was most frequent in patients with extremity cSCC (19/26 regional recurrences). The detail of failure patterns by anatomical site and surgical procedure is presented in Table 2.

**Table 2:**
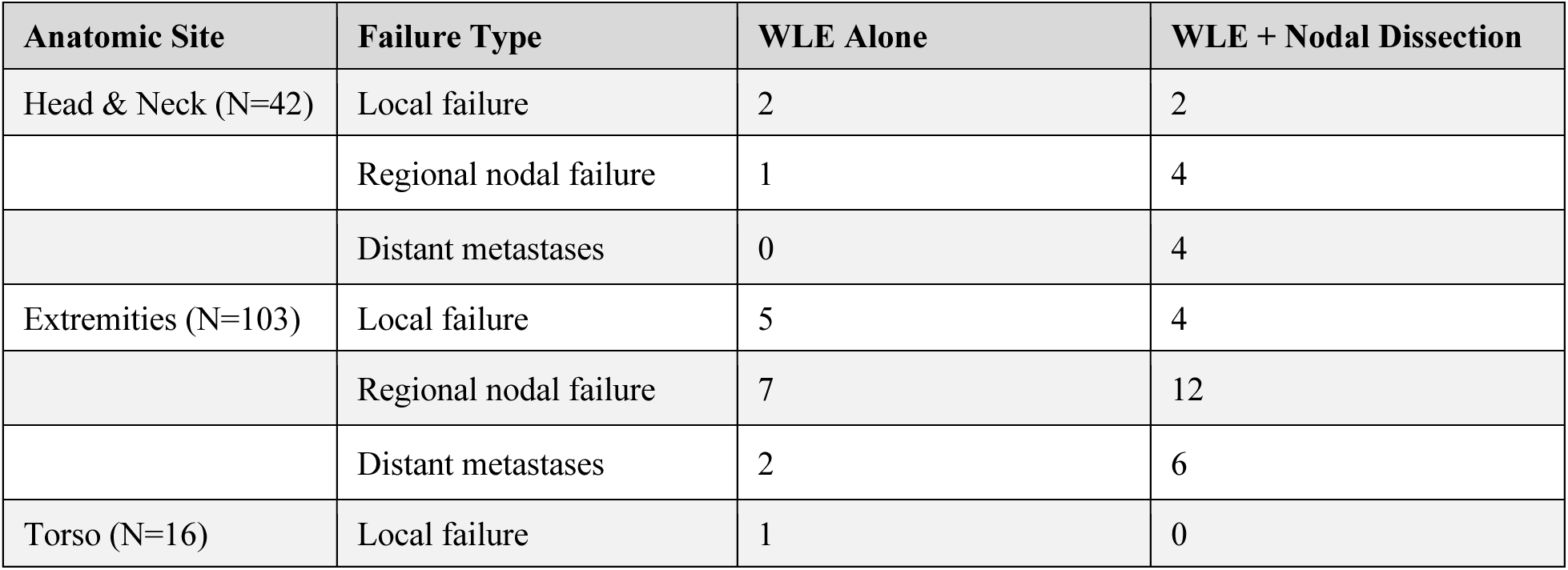
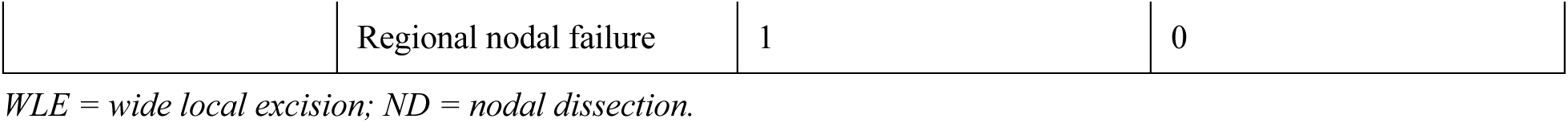
Patterns of failure by anatomic site and surgical procedure.

### BWH Staging and Failure Patterns

Tumour staging was also performed using the Brigham and Women Hospital (BWH) staging system (9,10) which classifies cSCC based on four risk factors: tumour diameter ≥2 cm, poorly differentiated histology, perineural invasion ≥0.1 mm and invasion beyond the subcutaneous fat (bone invasion automatically upgrades the tumour to BWH T3). BWH staging was applied to patients whose histopathological reports documented invasion of skin layer (N=135). The BWH T2b subgroup had the highest incidence of regional failure at 27.6% (13/47 patients), substantially exceeding rates in the T3 group (17.8%) and T2a group (6%). Every patient who failed regionally had at least one adverse pathological feature (either a positive/close margin or perineural invasion) and the majority were in the T2b category.

Perineural invasion was present in 27.7% of T2b patients and 18.5% of T3 patients. All patients with T2b or T3 stage BWH disease who experienced regional failure had PNI, with a statistically significant association (p<0.001). This reinforces the distinctly aggressive nature of the T2b subgroup in this population.

Cumulative incidence of local progression was observed most frequently with BWH T2a followed by T3 and T2b stages, while the highest cumulative incidence of regional progression was associated with BWH T3.

The median time to development of regional metastasis was 8.4 months.

## DISCUSSION

In this retrospective series of 161 surgically treated cSCC, regional recurrence (RR) was the most common mode of failure, with a 3-year cumulative incidence of 19% (95% CI: 13%–27%). It is higher than most of the western studies. (Table 4)

The current standard of care for cSCC consists of surgical excision of the primary tumour, with adjuvant radiotherapy for patients with high-risk features. Elective nodal dissection or sentinel lymph node biopsy (SLNB) is not currently recommended as standard practice. However, Ross et al. in their series reported SLNB positivity rates of 21 and 24% across anogenital and non-anogenital subtypes respectively, suggesting that nodal staging may benefit carefully selected high-risk patients.(11) Established risk factors for failure include advanced stage, positive or close margins, PNI, subcutaneous fat invasion, and poor differentiation.(10) Brantsch et al. reported a 4% risk of nodal metastasis and a 1.5% risk of disease-specific death in patients with head and neck cSCC. (12)

Our BWH staging results are concordant with those reported by Karia et al.(9,10) Patients in the T2b and T3 categories demonstrated higher rates of nodal recurrence, validating the prognostic utility of BWH staging. Notably, the T2b subgroup in our cohort had the highest regional failure rate (27.6%), exceeding that of T3 patients. This pattern likely reflects the combined burden of multiple moderate-risk factors in T2b disease, rather than any single dominant feature.

A critical limitation in the current classification criteria is the absence of a dedicated staging system for cSCC arising outside the head and neck region. The AJCC 7th edition did not differentiate by site; the 8th edition introduced a revised system exclusively for head and neck cSCC.(13,14) This leaves a significant gap for extremity and truncal primaries. Comparative analyses have demonstrated that the BWH system outperforms AJCC-7 in discriminating high-risk patients, Jambusaria et al. showed that in a cohort of 1808 patients, re-classification by BWH staging upstaged 86 tumours to T2b, with 70% of regional progression events occurring in this group.(15) Table 3 summarises key staging comparison studies. Consideration can be given to incorporate staging systems like BWH staging for stratification of patients with cSCC.

**Table 3:**
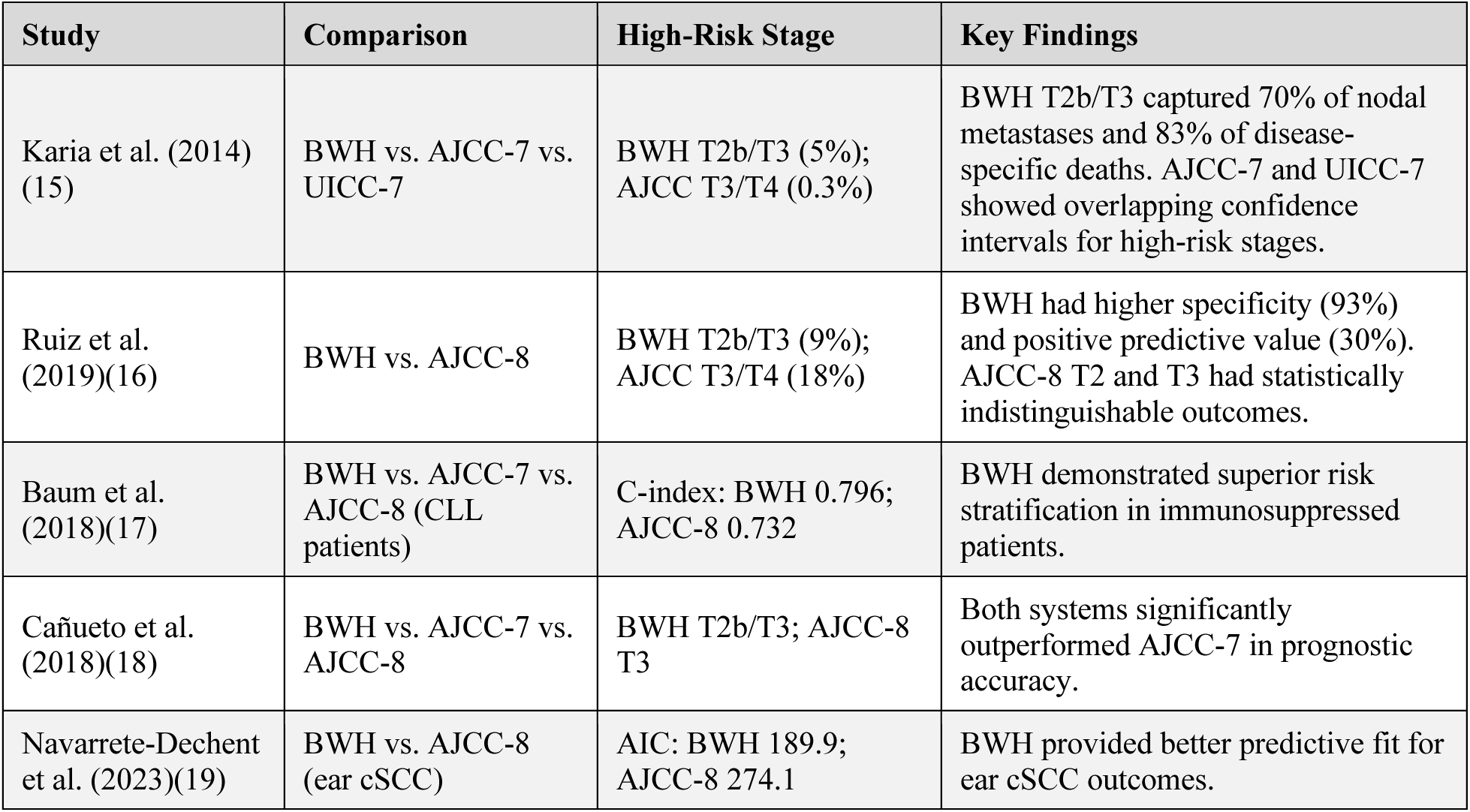
Comparison of cSCC staging systems across key studies.

The duration of follow up of these patients is not defined. The median time to regional metastasis was 8.4 months in our cohort, with most events occurring within the first year after surgery. Kraus et al. similarly reported a mean of 13 months to clinically significant nodal metastasis in head and neck cSCC.(20) Rowe et al. found that 69% of nodal metastases from cSCC of the skin, ear and lip were detected beyond 1 year, with over 90% within 3 years.(21) These data collectively suggest that the first 12 months following surgery with or without nodal dissection represents the high risk period, during which intensified clinical monitoring, including regular ultrasonography of the regional nodal basin should be considered.

Among reported series of extremity cSCC, regional recurrence rates in our cohort are at the higher end of the spectrum (Table 4). This may reflect the inclusion of Marjolin’s ulcers (n=21), a higher proportion of late presenting patients and the limited use of elective nodal staging.

**Table 4:** Comparison with published series of cSCC of the extremities.

| Study | N | Site | Follow-up | Regional Recurrence |
| --- | --- | --- | --- | --- |
| Joseph et al. (1992)(22) | 79 | Trunk/limbs | Mean 5.3 years | 7% (6/79 regional LN metastases) |
| Lifeso & Bull (1985)(23) | 112 | Extremities | 5–10 years | 6% LN metastasis (7/112) |
| Ames & Hickey (1980)(24) | Review | Extremities | Long-term | ~5% metastasis rate |
| Dormand et al. (2010)(25) | 104 | Upper/lower limbs | Median 5.2 years | 5% nodal metastasis (5/104) |
| Ribeiro et al. (2006)(26) | 112 | Lower limbs (Brazil) | 3 years | 4.5% regional (5/112) |
| Vazquez et al. (2008)(27) | 57 | Trunk/extremities (advanced) | Not specified (retrospective) | High frequency of LN metastasis (exact % not in abstract) |
| Nelson & Ashton (2017)(28) | 1,006 | Various incl. trunk/extremities | 5 years | <2% metastasis (trunk/extremities subset) |
| Fox et al. (2018)(29) | High-risk cohort | Extremities/trunk | Not specified | BWH T2b/T3 predicts >20% nodal risk |
| Huis et al. (2023) (30) | 103 (post-LND) | Various (metastatic cSCC) | Median 27 months | 33% overall; 35% nodal basin recurrence |
| Present study | 103 | Extremities | Median 2.4 years | 19% regional recurrence (3-year cumulative incidence) |

## LIMITATIONS

This study has several limitations inherent to its retrospective design. Patients were included at heterogeneous stages of disease and the number of patients is relatively small. Systematic assessment of sun exposure, occupational exposures, and immune status at presentation was not performed. Follow-up was variable and in some cases, incomplete; routine regional nodal imaging was not performed systematically during follow-up. Pathological reporting was not consistent across all cases, particularly regarding depth of invasion. Additionally, a subset of patients who were treated during the COVID-19 pandemic experienced prolonged gaps in follow-up, despite telephone outreach efforts.

## CONCLUSION

Regional recurrence is the predominant mode of failure which is higher than rates reported in most published Western series. Extremity cSCC and the BWH T2b pathological subgroup are associated with particularly high rates of regional failure. A comprehensive, staging system for cSCC is needed to guide risk stratification and management across all anatomical locations. These findings form the basis for prospective studies to better delineate predictors of regional failure and evaluate mitigating strategies, including elective nodal staging and consideration of post operative radiotherapy in appropriately selected high-risk patients.

## Data Availability

All data produced are available online at Pubmed

## Notes

### Competing Interest Statement

The authors have declared no competing interest.

### Author Declarations

The study used ONLY openly available human data that were originally located at: Tata Medical Center, Kolkata. The data that we used was aggregated/summary data.

